# Differential risk for COVID-19 in the first wave of the disease among migrants from several areas of the world living in Spain

**DOI:** 10.1101/2020.05.25.20112185

**Authors:** Carlos Guijarro, Elia Pérez-Fernández, Beatriz González-Piñeiro, Victoria Meléndez, Maria José Goyanes, Ma Esther Renilla, Maria Luisa Casas, Isabel Sastre, María Velasco, Alcorcon COVID Investigators

**Affiliations:** Internal Medicine Unit, Hospital Universitario Fundación Alcorcón. Alcorcón. Madrid. Spain; Infectious Diseases Section, Hospital Universitario Fundación Alcorcón. Alcorcón. Madrid. Spain; Research Unit, Hospital Universitario Fundación Alcorcón. Alcorcón. Madrid. Spain; Technology and Information Systems, Hospital Universitario Fundación Alcorcón. Alcorcón. Madrid. Spain; Microbiology Unit, Hospital Universitario Fundación Alcorcón. Alcorcón. Madrid. Spain; Emergency Unit, Hospital Universitario Fundación Alcorcón. Alcorcón. Madrid. Spain; Laboratory Unit, Hospital Universitario Fundación Alcorcón. Alcorcón. Madrid. Spain; Department of Medical Specialties and Preventive Medicine. Universidad Rey Juan Carlos. Alcorcón. Madrid. Spain; Department of Social, Senior and Public Health Services. City Council. Alcorcón

**Author notes:** **Correspondence to.** Carlos Guijarro, Internal Medicine Unit. Hospital Universitario Fundación Alcorcon - Universidad Rey Juan Carlos. Budapest 1, 28922 Alcorcón, Madrid (Spain), **e-mail:**. **Alcorcón COVID Investigators: (Collaborators):** Alejandro Algora, Juan Carlos Alonso-Punter, Gregorio Bonilla Zafra, Mercedes Bueno-Campaña, Virgilio Castilla, Ana Isabel Diaz-Cuasante, Aurora Fabero, Rosa Maria Fariña, Isabel Ma Galán, Isabel González-Anglada, Ma Mercedes Izquierdo, Susana Lorenzo, Juan Emilio Losa, Margarita Mosquera, Carmen Noguera, Montserrat Pérez-Encinas, Gil Rodríguez-Caravaca, José Francisco Valverde.

## Abstract

**Introduction and Objectives:** Little is known regarding the relevance of racial / ethnic background on the risk of COVID-10 infection, particularly in Europe. We evaluated the risk for COVID-19 among migrants from different areas of the world within a context of universal free access to medical care.

**Materials and Methods:** We conducted a population-based cohort analysis of the cumulative incidence of PCR-confirmed COVID-19 among adult residents at Alcorcón (Spain) in the first wave of the disease up to April 25, 2020.

**Results:** The crude cumulative incidence among migrants (n=20419) was higher than among Spaniards (n=131599): 8.81 and 6.51 and per 1000 inhabitants respectively (p<0.001) but differed by world region of origin. By negative binomial regression, adjusted for age and sex, relative risks (RR) for COVID-19 were not significantly different from Spaniards for individuals from Europe, Asia or Northern Africa. In contrast, there was a marked increased risk for Sub-Saharan Africa (RR 3.66, 95% confidence interval (CI) 1.42-9.41, p=0.007), Caribbean (RR 6.35, 95% CI 3.83-10.55, p<0.001) and Latin-America (RR 6.92, 95% CI 4.49-10.67, p <0.001).

**Conclusions:** Migrants from Sub-Saharan Africa, the Caribbean and Latin-America, as opposed to Spaniards or migrants from Europe, Northern Africa or Asia exhibited an increased risk for COVID-19. Our data suggest a role for ethnic background in the risk for COVID-19. Migrants from some areas of the world may deserve a closer attention both for clinical and epidemiological reasons.

## Resumen

**Introducción y Objetivos:** Existen pocos estudios sobre el potencial papel de los orígenes raciales / étnicos en el riesgo de infección COVID-10, particularmente en Europa. Evaluamos el riesgo de COVID-19 entre los migrantes de diferentes zonas del mundo en un contexto de acceso universal gratuito a la atención médica.

**Materiales y Métodos:** Realizamos un análisis de cohortes poblacional de la incidencia acumulada de COVID-19 confirmado mediante PCR entre los residentes adultos en Alcorcón (España) en la primera oleada de la enfermedad hasta el 25 de abril de 2020.

**Resultados:** La incidencia acumulada bruta entre los migrantes (n=20419) fue mayor que entre los españoles (n=131599): 8,81 y 6,51 y por cada 1000 habitantes respectivamente (p<0,001), pero difería según la región de origen mundial. Mediante regresión binomial negativa, ajustada por edad y sexo, los riesgos relativos (RR) para COVID-19 no fueron significativamente diferentes de los españoles para individuos provenientes Europa, Asia o el norte de África. Por el contrario, hubo un marcado aumento del riesgo para el África subsahariana (RR 3,66, intervalo de confianza del 95% (CI) 1,42-9,41, p=0,007), Caribe (RR 6.35, 95% CI 3.83-10.55, p<0,001) y Latinoamérica (RR 6.92, 95% CI 4.49-10.67, p <0,001).

**Conclusiones:** Los migrantes procedentes del África subsahariana, el Caribe y América Latina, a diferencia de los españoles o migrantes procedentes de Europa, el norte de África o Asia, presentaron un mayor riesgo para COVID-19. Nuestros datos sugieren un papel para el origen étnico en el riesgo de COVID-19. Los migrantes de algunas zonas del mundo pueden merecer una atención más cercana tanto por razones clínicas como epidemiológicas.

## Introduction

COVID-19 pandemic is imposing a tremendous challenge to the humankind with dramatic health and economic consequences [1–3]. Recent reports have highlighted an increased COVID-19 related mortality among migrants and ethnical minorities in Western countries [4–10]. However, the clinical outcomes (in particular mortality) associated to ethnic minorities are inextricably connected to different socioeconomic backgrounds, pre-existing medical conditions, and unequal access to medical services [11–14]. There have been a number of mass-media reports and calls to action but very few sounded scientific publications addressing this issue, most of them from the USA or United Kingdom, and are virtually absent from other European countries [15–18].

We sought to describe the incidence of COVID-19 in the first wave of the disease among Spaniards and migrants from different areas of the world living in Alcorcón, a city in the suburbs of Madrid (Spain) with a substantial proportion of foreign residents. The differential proportion of migrants into Spain from several areas of the world may provide an opportunity to evaluate COVID risk in these populations.

## Methods

Population based cohort study conducted at Alcorcón (Madrid), with a total population of about 170,000 inhabitants. The population at risk was defined as all adults included in the official municipal live registry of population of the City Council of Alcorcón (last updated March 14, 2020).[19] Distribution of the population by nationality and country of origin stratified by age and sex was obtained from this registry.

Case was defined as a patient with a COVID-19 diagnosis at Hospital Universitario Fundación Alcorcón confirmed by Reverse Transcription (RT)-Polymerase Chain Reaction (PCR) for SARS-CoV2 for adults residing in Alcorcón. Incident cases of COVID-19 were obtained from the Electronic Patient Record (Selene ©). COVID-19 clinical diagnosis was established by the attending clinicians at the Emergency Room according to European Center for Disease Control and Prevention (ECDCP)-World Health Organization Criteria (WHO) [20].

For molecular diagnosis of SARS-CoV-2 infection, nasopharyngeal swabs from symptomatic patients were processed by automatized extraction using the MagNaPureLc instrument (Roche Applied Science, Mannheim, Germany) and real time reverse-transcription PCR using the SARS-Cov-2 nucleic acid detection Viasure kit (CerTestBiotec S.L.), following the manufacturer’s instructions. For this rRT-PCR, we used Bio-Rad CFX96™ Real-Time PCR Detection System. We amplified two different viral regions: ORF1ab gene (FAM channel), N gene (ROX channel), and the internal (HEX channel). Cycle threshold values ≤40 were considered positive. Positive and negative controls were included in each run for every assay.

An anonymized set of data was extracted from the electronic patient record (from February 1st to April 25th) containing the following parameters: age, sex, nationality, country of birth, city of residence, date of COVID-19 diagnosis (clinical), date and results of SARS-CoV2 PCR, clinical evolution / outcomes (hospital admission, critical care admission, hospital discharge, length of hospital stay, and in-hospital death). For the present study patients whose residence was outside the city of Alcorcón were excluded.

Patients were classified according to their country of origin into one the following groups: a) Spain; b) European Union (including Switzerland and Norway); c). Eastern Europe (including Russia); d) Asia, e). Australia & New Zealand; f) Northern Africa; g) Sub Saharan Africa; h) Latin America (Mexico and continental Latin-American countries); i) Caribbean; j) United States of America & Canada.

Statistics. Results are described as means (± standard deviation), medians (interquartile range) or percentages as appropriate. Quantitative variables were compared by the student’s t test, ANOVA, or U Mann-Whitney’s test, as appropriate. Qualitative variables were compared by the Chi^2^ test or the Fisher’s exact test as appropriate.

Aggregated data by world zone (or country), sex and age group (<30, 30-39, 40-49, 50-59, 60-69, 70-79, 80-89 and >=90 years) were analyzed. Incidence rates and 95% exact Poisson confidence intervals were calculated by world zone of origin. Multivariate negative binomial regression model with robust variance was used to estimate the incidence rate by world zone adjusted for sex and age. These models are appropriated to analyze over-dispersed count data [21,22]. Statistical significance was assumed for p values <0.05. All statistical analyses were conducted with Stata 14 (StataCorp LLC, Texas, USA).

The protocol received the approval of the local Independent Review Board / CEIm (Medicines Research Ethical Committee).

## Results

Alcorcon city registry (last updated on March 14) comprised 152,018 individuals 17 years of age or older [19]. Among them, 20,419 (13.4%) residents had a nationality different from Spain. Their countries of origin (and associated areas of the world) are summarized in Table 1. Different areas of the world were chosen because of geographical, socioeconomic and ethnic background reasons provided there was a minimum of 500 individuals per area. Migrants from the USA, Canada, Australia and New Zealand) added up less than 200 residents and were removed for further analysis.

Migrant residents were substantially younger than Spaniards (median age 40 years IQR 30-50 vs 49,5 years IQR 38-67 respectively, Supplemental Figure 1), with a slightly higher proportion of males (48,7 vs 47,6% p=0,002). Indeed, age distribution pyramids in the reference population were strikingly different for Spaniards and migrants (Supplemental Figure 2).

A total of 1691 residents in Alcorcon were diagnosed as having COVID-19 following ECDCP-WHO clinical criteria [20] from February 1 until April 25 at our hospital (Figure 1A). The number of cases increased steadily until the end of March and declined thereafter (Figure 1B), roughly 2 weeks after the lockout Decree of the Spanish Government (March 14, 2020)[23]. In order to avoid potential misdiagnosis, we restricted our analysis to the 1036 cases with a positive PCR for SARS-CoV2 sample (hereafter COVID-19-PCR+ cases). Median age of COVID-19-PCR+ patients was 71 years (IQ range 54-79), and 57.2 % were male.

**Figure 1.**
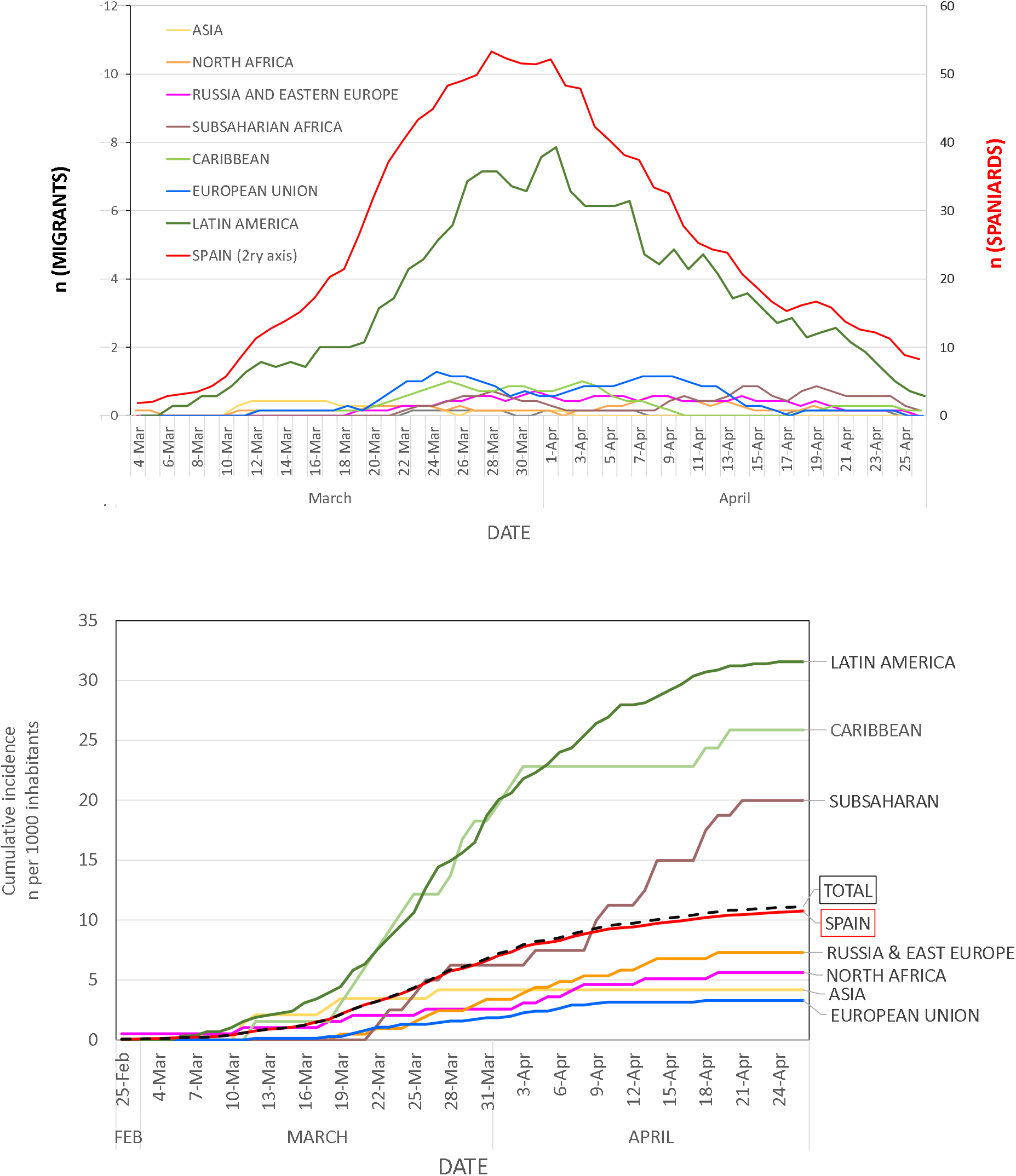
Evolution of clinical diagnosis of COVID-19. Panel A: daily diagnosis (graph represents for every date the 7 day moving average = number of daily diagnosis of the preceding week) at Hospital Universitario Fundación Alcorcon. Panel B Cumulative incidence rate of new COVID-19 clinical diagnosis per region of the world at Hospital Universitario Fundación Alcorcón.

The global accumulated COVID-19-PCR+ incidence was 6.81 per 1000 inhabitants. The global accumulated COVID-19-PCR+ incidence in Spaniards was 6.50 cases per 1000 inhabitants as compared to 8.82 per 1000 for non-Spaniards (p<0.001; Table 2, Figure 1B). Spanish COVID-19-PCR+ cases were substantially older than migrants (median age 73 years, IQR 62-80 vs 51,5, IQR 41,7-58, p<0,001), with a non-significant higher proportion of males (58.6% vs 52.5%; p=0.15) (Supplemental figure 3). The global cumulative incidence increased dramatically with age for both Spaniards and migrants (Supplemental Figure 3A).

We next evaluated cumulative incidence rates of COVID-19-PCR+ for natives from different regions of the world as compared to Spaniards (Table2, Figure 1B). Unadjusted incident rates for individuals from Europe, Asia and North Africa were lower than average, while for Sub-Saharan Africa, Caribbean and Latin-America were higher. Cumulative incident rates for most representative countries are described in Supplemental Table 1.

In order to have a better understanding of risks for Spaniards and migrants from different regions of the world we conducted a multivariate negative binomial regression analysis [21,22] adjusting for age and sex, as they are proven determinants of COVID-19 risk (Figure 2). The adjusted relative risks for COVID-PCR+ among migrants from Asia, European Union and Eastern Europe did not significantly differ from Spaniards (Table 3, Figure 2). In contrast, the relative risk for Latin-America migrants was about 7-fold higher than that for Spaniards (RR 6.92; 95% CI 4.49-10.67, p <0.001). In addition, the adjusted risk was also increased for Sub-Saharan Africa (RR 3.66 95% 1.42-9.41, p=0.007) as well as Caribbean (RR 6.35 95% CI 3.83-10.55, p<0.001) in a highly clinical and statistically significant manner.

**Figure 2.**
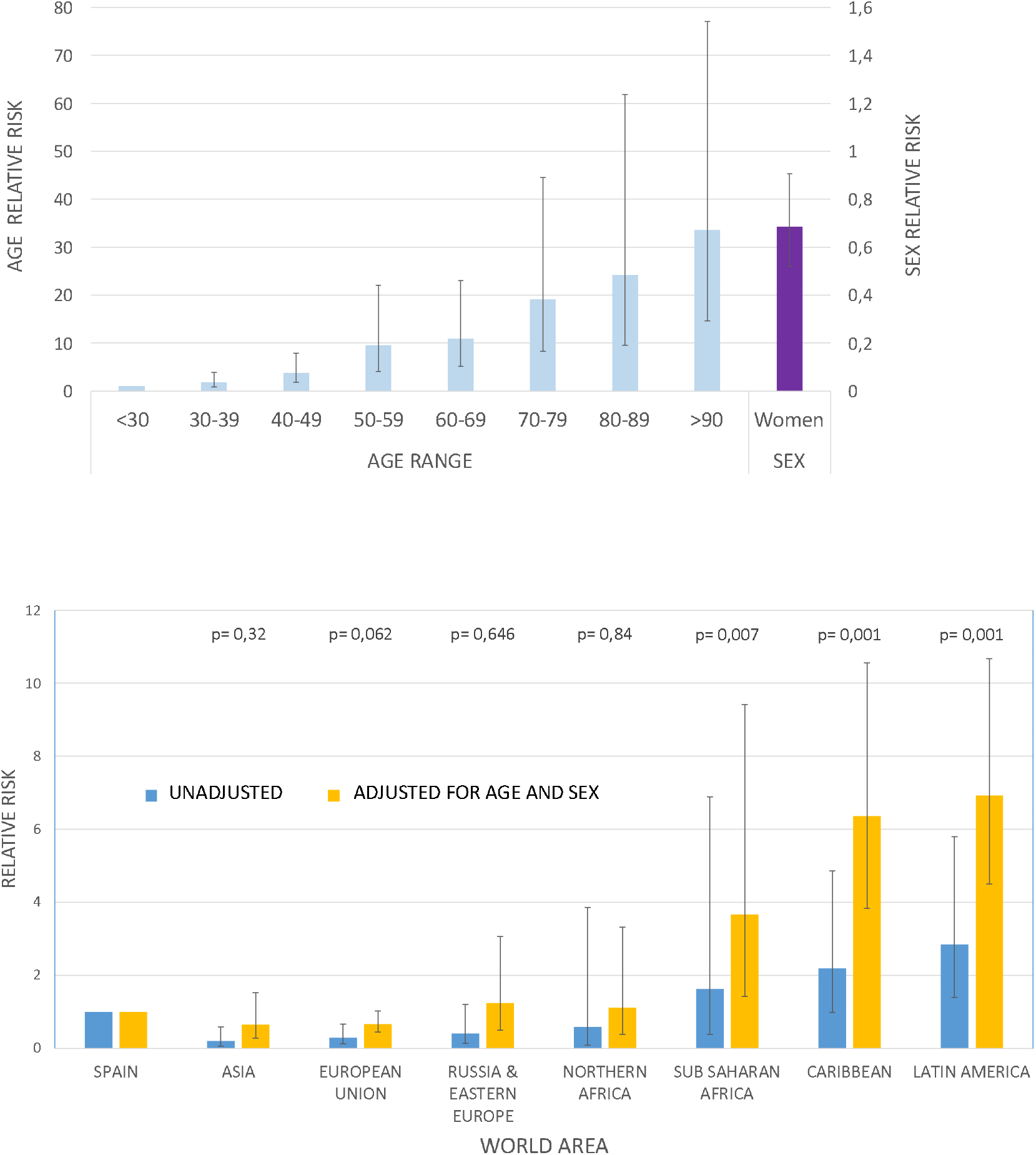
Relative risks for COVID-19 for age intervals (reference age < 30 years), sex (Panel A), as well as areas of the world (reference Spain) calculated by negative binomial regression analysis (Panel B).

As a sensitive analysis (Table 3), we repeated our estimation with wider criteria (clinical diagnosis regardless of PCR results; n=1,691) and stricter criteria (patients requiring hospital admission because of moderate-to-severe COVID-19, n=877). In both cases results remained essentially unchanged. Adjusted risks for Sub-Saharan Africans, Caribbeans and Latinos were about 3, 6 and 7 fold higher in all clinical scenarios (Table 3). As an exploratory evaluation, we performed a similar analysis for countries that had at least 800 inhabitants in Alcorcón (Table 1). Citizens from Romania, Ukraine, China, or Morocco did not exhibit increased rates for COVID-19 cumulative incidence (Supplemental Figure 4). In agreement with regional data, migrants from all Latin American countries showed a consistent increased risk for COVID-19.

Severe COVID-19, defined as death, critical care admission or hospital stay longer than 7 days occurred in 63% of the Spaniards as compared to 33% for migrants. Similarly, unadjusted mortality was higher for Spaniards (25% of admissions) than for migrants (6%). Most likely this mirrors the advanced age of patients with severe disease (median 76 years IQR 69-82 years) or death (median 79 years IQR 74-85), an age range virtually absent in migrants (Supplemental Figures 1, 2). Our data do not support a more severe disease among migrants residing in Spain.

## Discussion

On a population-based approach, our data highlight that the incidence of COVID-19 varies among migrants from different areas of the world as well as compared to the Spanish reference population. A major finding of our work is an apparent higher risk for COVID-19 for individuals from Sub-Saharan, Caribbean or Latin-American origin, as compared to migrants from Europe, Asia or Northern Africa or Spaniards. Preliminary data from other area in Madrid are consistent with our findings [10]. The higher risk is consistently detected when it is estimated globally for their regions of origin or for particular countries. Furthermore, the risk is clearly enhanced when adjusted for age and gender. In addition, we found a lower unadjusted relative risk for COVID-19 risk for other European citizens as compared to Spaniards. However, migrants represent a younger population when compared to Spaniards [24,25], as it is the case in our setting. Indeed, when adjusted for age and gender the relative risk for COVID-19 for migrants from Europe (both for European Union and Eastern Europe) did not differ from Spaniards. Similarly, the adjusted incidence rate of COVID-19 in migrants from Asia or Northern Africa was not different from the reference population.

The reasons for this apparent risk for COVID-19 among migrants from different areas of the world are not apparent. First, all migrants have equal access to the virtually universal health coverage available for Spaniards. Second, most migrants have low-wage jobs and pertain to a low socioeconomic status [26,27],. This is also the case for ‘European’ migrants since this population is vastly represented by Romania and Ukraine in our city. An increased prevalence of hypertension or obesity may underlie a higher risk for migrants from Sub-Saharan Africa or Latin America, as it has been described in blacks or Hispanics in the USA [28]. However, in our setting migrants from these areas do not seem to exhibit higher prevalence of obesity, hypertension, diabetes or cardiovascular disorders. This may be related to a younger age and the so called ‘healthy migrant effect’ [24,25,29,30].

One strength of our study is the population-based approach. We used the best available updated registry of citizens residing in the reference population. This is also likely the case for migrants, since a number of social services rely upon the actual place of residence [19]. Our estimation of the incidence of COVID-19 is a reasonable approach since the population primarily attended by our hospital is precisely the city of Alcorcón and there is no other public hospital in the city [31]. In case of an emergency such as COVID-19, it is likely that citizens rely even more into their local facilities. In any event, it is almost certain that a number, albeit small, of Alcorcón residents have been attended because of COVID-19 in hospitals different from ours and thus our data are likely an underestimation of the real incidence. We have excluded from our analysis 16% of COVID-19 cases attended in our hospital whose residence was not Alcorcón. Official data from the Community of Madrid indicates that our hospital receives a slightly higher number of patients from other areas than viceversa [31]. Accordingly, our underestimation of COVID-19 incidence in Alcorcón may be somewhere between 10-15%. We do not believe that these data modify substantially the relative incidence for migrants, since the proportion of COVID-19 migrants excluded from our study because of residence outside Alcorcon was essentially identical (17%) as that for Spaniards (16%).

Another critical issue relates to the actual access to medical care for migrants. Since July 2018 all individuals resident in Spain have access to free medical care in conditions virtually identical to Spaniards [32]. It might be argued that some migrants may rely less on primary care physicians and thus show a disproportionate use of hospital services [25,33]. However, our sensitive analysis, restricted to more severe COVID-19 requiring hospital admission does not support this potential overuse of hospital resources.

Some limitations of our study must be addressed. First, we did not have local data regarding socioeconomic status, education or other health conditions for migrants. However, on aggregate, migrants from Romania, Ukraine or Morocco residing in Spain belong to a relatively lower socioeconomic stratum all along Spain [27] and do not exhibit an increased risk for COVID-19 as opposed to migrants from Sub-Saharan Africa or Latin-America, suggesting that socioeconomic status may not be a major reason for the differences in COVID-19 incidence.

Once the above-mentioned factors do not seem to adequately explain this increased incidence, we should pay attention to the ethnic background of the populations. Interestingly, a recent report from a large database from the Veteran administration suggests that Black and Hispanic individuals in the USA are experiencing an excess burden of Covid-19 not entirely explained by underlying medical conditions [28], in agreement with our results. Interestingly, recent reports from the USA suggest that the increased risk for black minorities does not occur in Asian minorities, also in agreement with our data [34]. These data point to a different ethnic background, in addition to other socio-economic reasons, as a potential risk factor for COVID-19.

There are limited data regarding different susceptibility to respiratory viruses from different ethnic backgrounds. Some studies have suggested a greater susceptibility of racial minorities to influenza [35]. However, many studies have failed to dissociate clinical outcomes from socioeconomic issues, particularly unequal access to health care. Different susceptibilities to COVID-19 with a genetic background with unequal world distribution are just beginning to be explored [36–38].

Finally, our data do not suggest an increased mortality risk for migrants. Of note, black and Hispanic individuals did not exhibit increased mortality in the Veteran Administration database [28], in contrast with other reports in the USA [6]. These data suggest that health care access, rather than ethnicity may be the major determinant for survival in COVID-19.

In summary, we report a selective increased risk for COVID-19 among certain migrant populations in Spain: Sub-Saharan, Caribbean and Latin-America that is not related to unequal access to health care. These groups may deserve a particular attention, particularly when our country, as well as others, is experiencing an increase in COVID-19 incidence following the de-escalation of previous severe social distancing measures. In addition, our data are compatible with different genetic susceptibility to COVID-19 among diverse ethnic backgrounds that warrants further studies. Regardless of the reasons underlying the differential risk of COVID-19 in different migrant populations, our data can help to identify individuals or groups at particularly high risk who may merit special attention for both clinical and epidemiological reasons.

## Data Availability

No individual patient data are avaiulable

## Acknowledgements

There was no funding for the present work

We are grateful to Dr F. Rodriguez-Artalejo for critically reviewing the manuscript.

## Notes

### Competing Interest Statement

The authors have declared no competing interest.

### Funding Statement

No funding was available for this manuscript

### Author Declarations

IRB Hospital Universitario Fundacion Alcorcon

### Summary of Updates

Little is known regarding the relevance of racial / ethnic background on the risk of COVID-10 infection, particularly in Europe. We evaluated the risk for COVID-19 among migrants from different areas of the world within a context of universal free access to medical care. We conducted a population-based cohort analysis of the cumulative incidence of PCR-confirmed COVID-19 among adult residents at Alcorcon (Spain) in the first wave of the disease. The crude cumulative incidence among migrants (n=20419) was higher than among Spaniards (n=131599): 8.81 and 6.51 and per 1000 inhabitants respectively (p<0.001) but differed by world region of origin. By negative binomial regression, adjusted for age and sex, relative risks (RR) for COVID-19 were not significantly different from Spaniards for individuals from Europe, Asia or Northern Africa. In contrast, there was a marked increased risk for Sub-Saharan Africa (RR 3.66, 95% confidence interval (CI) 1.42-9.41, p=0.007), Caribbean (RR 6.35, 95% CI 3.83-10.55, p<0.001) and Latin-America (RR 6.92, 95% CI 4.49-10.67, p <0.001). Migrants from these areas of the world may deserve a closer attention both for clinical and epidemiological reasons.

