## Supplemental Table for "Differential risk for COVID-19 in the first wave of the disease among migrants from several areas of the world living in Spain"

| Supplemental Table 1. COVID-19 PCR+ patients at Alcorcón. Accumulated incidence at Alcorcón by representative countries (February-April 2020) | | | | | | | | | |
| --- | --- | --- | --- | --- | --- | --- | --- | --- | --- |
|  |  |  | **Europe** | | **Asia** | **Northern Africa** | **Latin-America** | | |
|  | **All** | **Spain** | **Romania** | **Ukraine** | **China** | **Morocco** | **Venezuela** | **Colombia** | **Peru** |
| Cases (no) | 943 | 856 | 7 | 7 | 2 | 6 | 8 | 19 | 38 |
| Age (median) | 72.0 | 73.0 | 42.0 | 58.0 | 46.0 | 70.5 | 55.0 | 46.0 | 52.0 |
| Interquartile range | 59-79 | 62-80 | 38-55 | 57-62 | 44-48 | 44,7-75 | 53-67 | 38-52 | 38-58 |
| Male (%) | 55.0 | 58.6 | 57,1 | 0.0 | 50.0 | 66.7 | 62,5 | 47.4 | 47.0 |
| Accumulated Incidence | 6.54 | 6.08 | 1.51 | 3.75 | 2.30 | 3.18 | 7.94 | 16.13 | 36.05 |
| Poisson Exact 95% confidence interval | 6.13-6.98 | 6.08-  6.96 | 0.61-  3.10 | 1.51-  7.73 | 0.28-  8.31 | 1.17-  6.93 | 3.43-  15.65 | 9.71-  25.19 | 25.51-49.49 |
