## Supplemental figures for "Differential risk for COVID-19 in the first wave of the disease among migrants from several areas of the world living in Spain"

Supplelmental Figure 1. Age group distribution of Spaniards and Migrants living in Alcorcon (March 14, 2020)

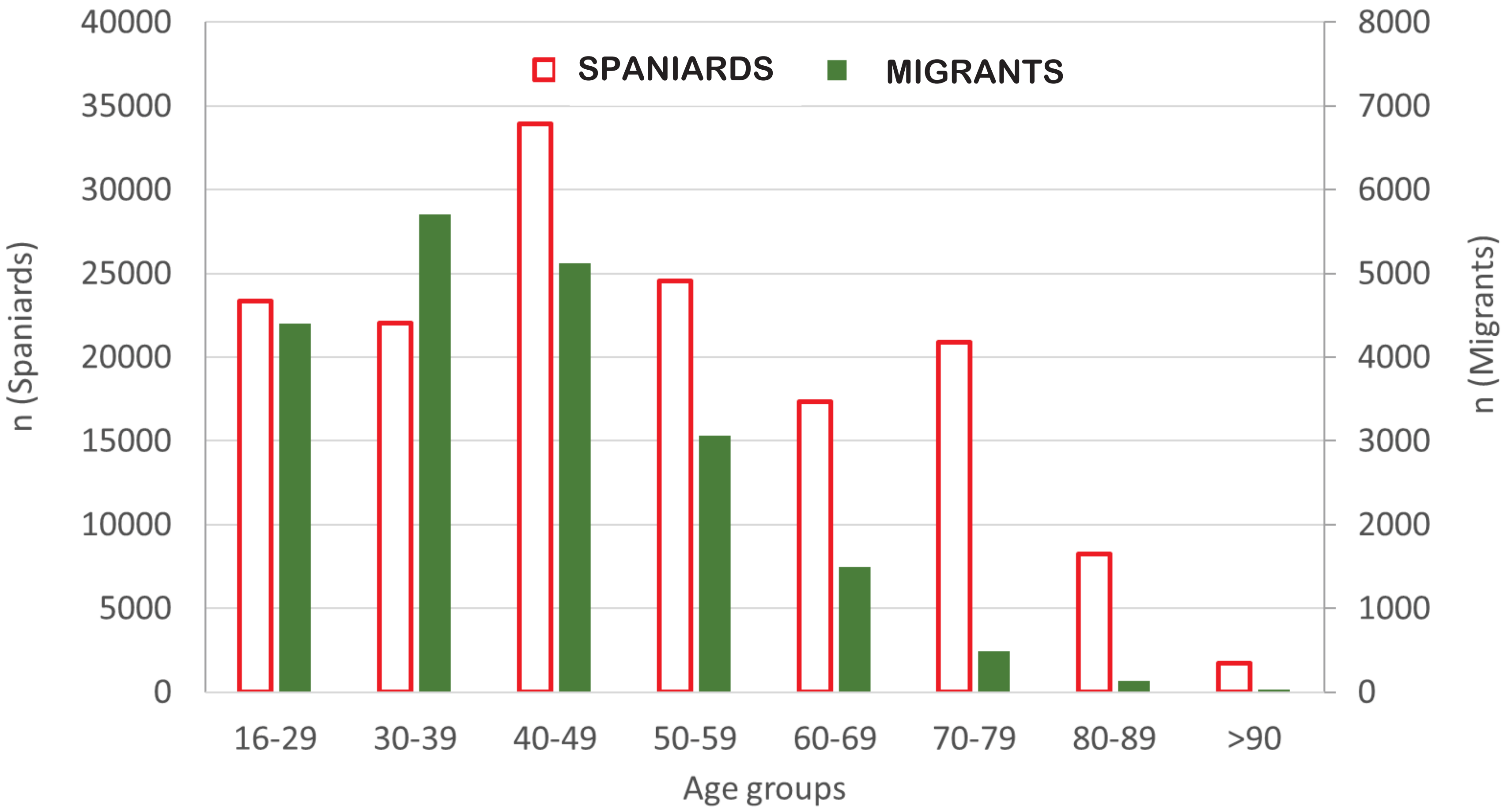

Supplemental Figure 2. Age pyramid distribution of Spaniards and Migrants from different regions of the world living in Alcorcon (March 14, 2020)

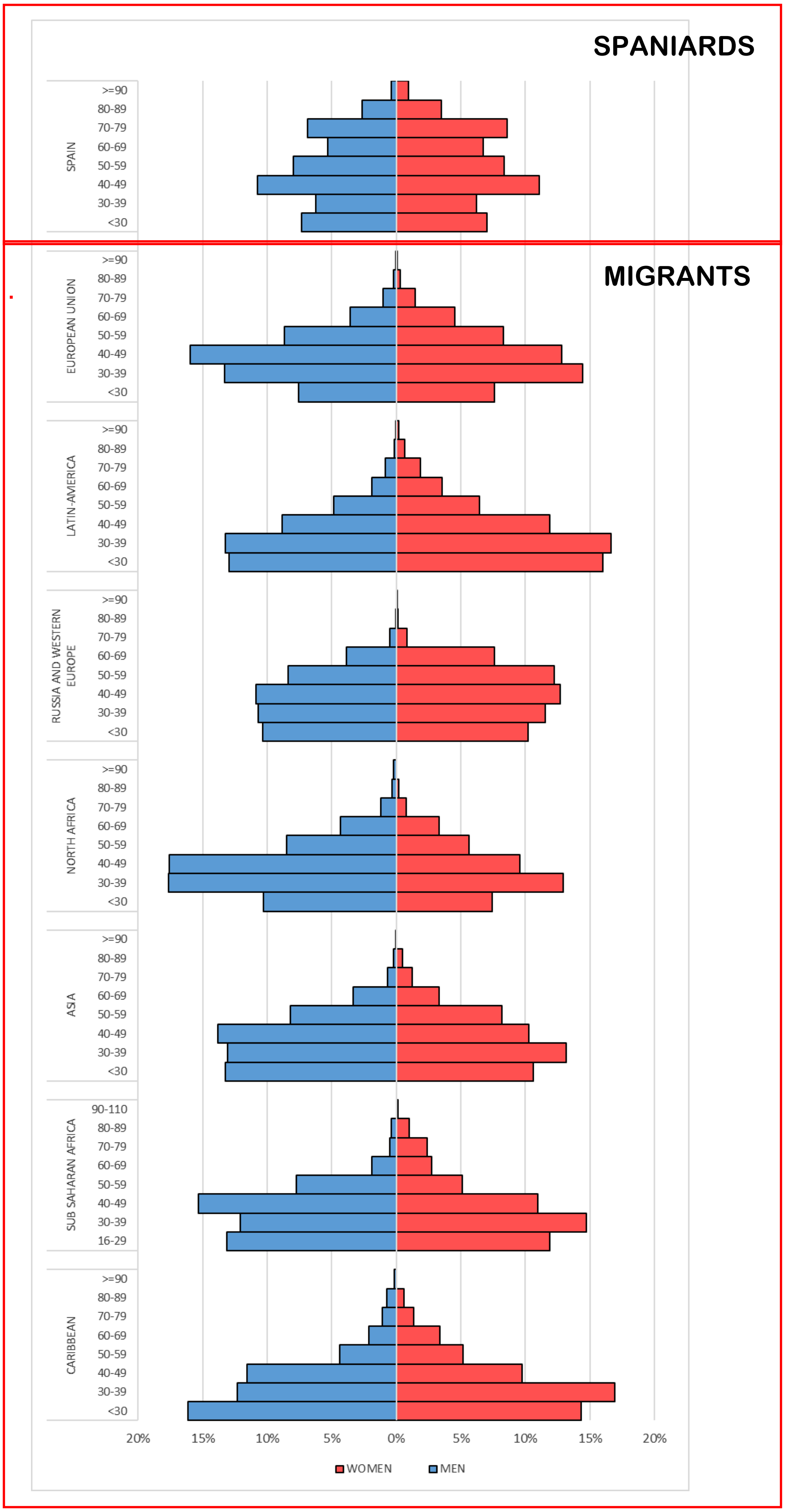

Supplemental Figure 3. Age group distribution of global population and patients diagnosed as having RT-PCR confirmed COVID 19 in Alcorcón until April 25. Upper panel: global population; Middle panel Spaniards; Lower Pannel Migrants-

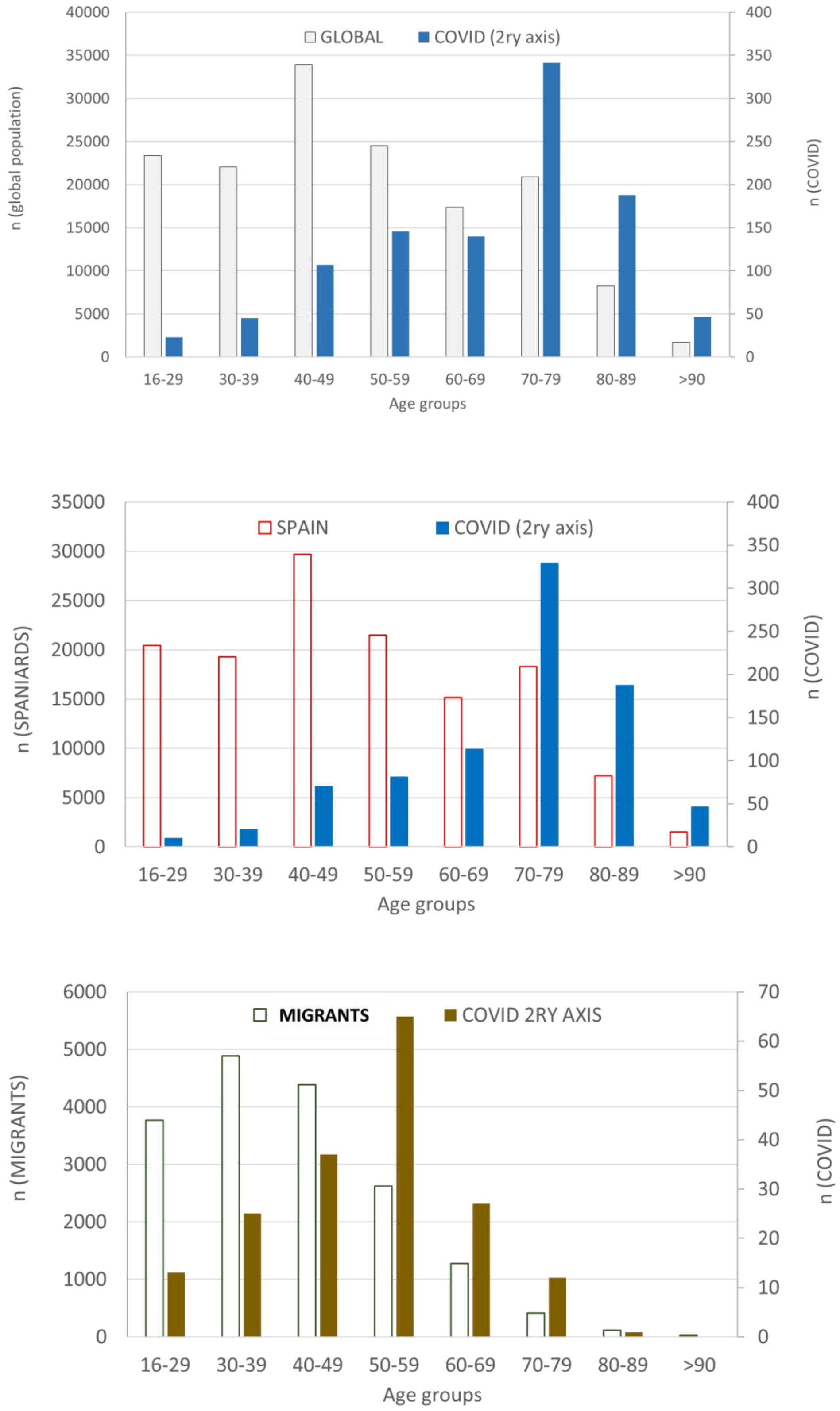

Supplemental Figure 4- Relative risks and 95% confidence intervals for RT- PCR-confirmed COVID-19 in migrants from countries with at least 1000 inhabitants in Alcorcon (reference Spain=1) calculated by negative binomial regression analysis. P values are depicted for the adjusted risks

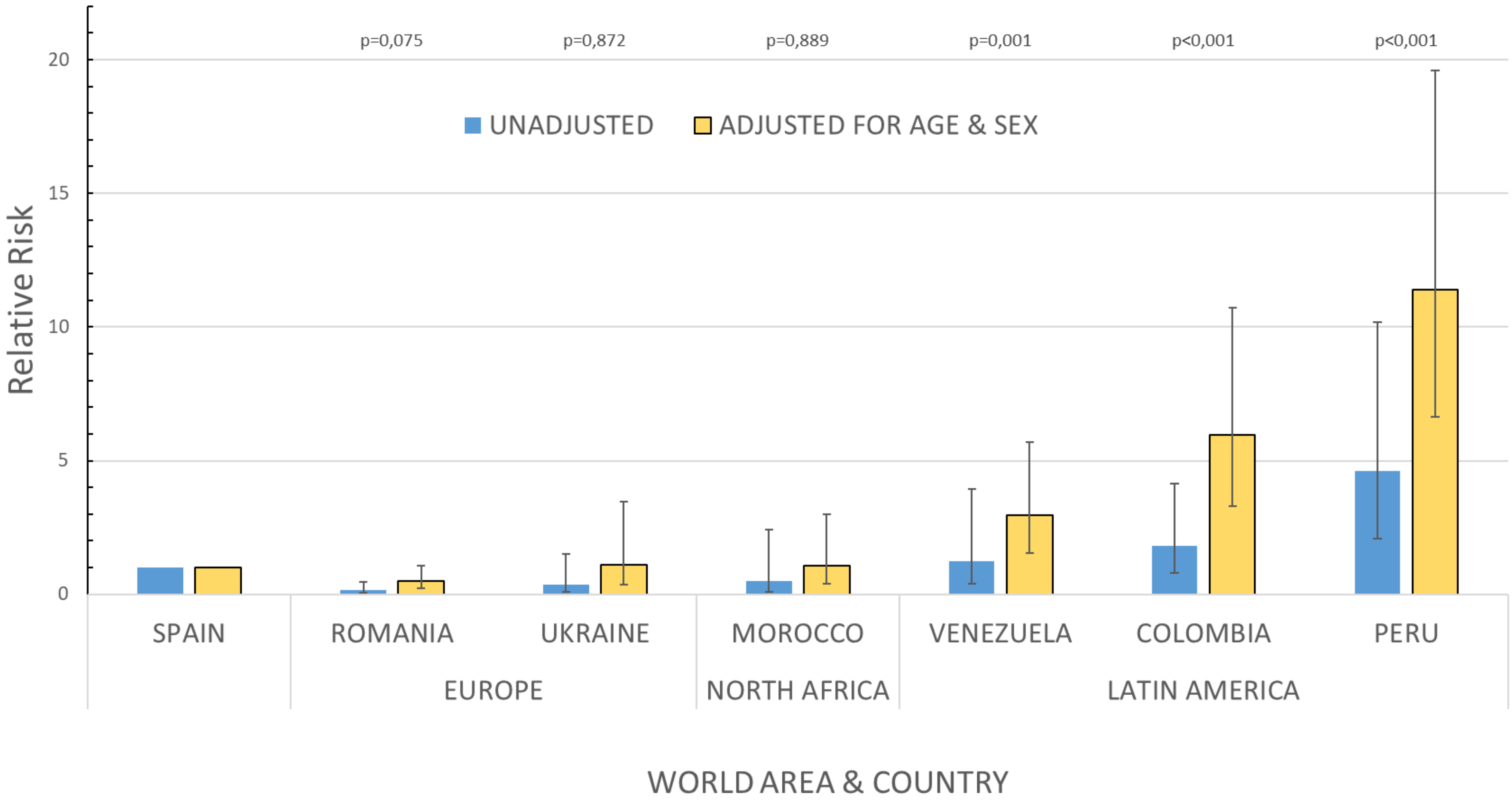
